# Statistical Analysis Plan: The Effect of Automated External Defibrillators for Cardiac Arrests in Private Homes An Observational Study

**DOI:** 10.1101/2023.08.01.23293407

**Authors:** Lars W. Andersen, Mathias J. Holmberg, Asger Granfeldt, Nikola Stankovic, Maria Høybye, Fredrik Folke, Lisa Caulley

## Abstract

This document describes the statistical analysis plan for the study “The Effect of Automated External Defibrillators for Cardiac Arrests in Private Homes”. This will be an observational study using prospectively collected data from the CARES registry in the United States. We will include patients with an out-of-hospital cardiac arrest in a private home. The exposure will be application of an AED and the primary outcome will be survival to hospital discharge. Assuming that there is no causal effect of AED application in those with a non-shockable rhythm, we will use a “difference-in-difference” approach to estimate the causal effect of AED application in those with a shockable rhythm.

## PREFACE

This document describes the statistical analysis plan for the study “The Effect of Automated External Defibrillators for Cardiac Arrests in Private Homes”. The statistical analysis plan will be finalized and published online prior to receipt of data. Any discrepancies between the statistical analysis plan and the final manuscript will be clearly described in the manuscript and a rationale for any changes will be provided.

## BACKGROUND

We have previously shown, using data from the Cardiac Arrest Registry to Enhance Survival (CARES) registry, that application of an automated external defibrillator (AED) is associated with improved outcomes in patients with shockable out-of-hospital cardiac arrest in a public location.[1] Specifically, in a propensity matched cohort, we found that survival was improved from 42% to 60% when the patient had an AED applied and had a shockable rhythm. There was no association between AED application and outcomes for those who had a non-shockable rhythm consistent with the mechanism of effect (i.e., defibrillation).[1] We subsequently did a cost-effectiveness analysis of public AEDs finding that these were cost-effective under many circumstances.[2] These findings have together helped inform decision-making and guidelines. However, there is an unanswered question remaining, namely whether AEDs in private homes (where approximately 70% of cardiac arrests occur) are effective and cost-effective.

## MAIN RESEARCH AIM

To estimate the effect on survival to hospital discharge of application of an AED, as compared to no AED, in patients with cardiac arrest and a shockable rhythm in a private home.

## GENERAL METHODS

### Overview

This will be an observational study using prospectively collected data from the CARES registry in the United States. We will include patients with an out-of-hospital cardiac arrest in a private home. The exposure will be application of an AED and the primary outcome will be survival to hospital discharge. Assuming that there is no causal effect of AED application in those with a non-shockable rhythm, we will use a “difference-in-difference” approach to estimate the causal effect of AED application in those with a shockable rhythm.

### Data source

We will be using data from the CARES registry from 2013 to 2022. CARES is a United States-based registry of non-traumatic out-of-hospital cardiac arrest. Cardiac arrests are included if resuscitation is attempted. This is defined as either 1) resuscitation performed by first responders or Emergency Medical Services (EMS), or 2) defibrillation prior to arrival of first responders or EMS. Cases with resuscitation terminated due to “do not resuscitate” directives or obvious signs of death are not included. Neither are cardiac arrests that did not involve 911 dispatch, e.g., during intrahospital transport.

The registry has a catchment area of approximately 175 million people. Multiple data sources, including data from dispatch centers, 911-responders, and hospitals are used to create a single record for each cardiac arrest event. Additional information is provided on the CARES website.[3]

### Patients

We will include patients with non-traumatic out-of-hospital cardiac arrest recorded in the CARES registry. We will only include patients aged ≥ 1 year with a cardiac arrest in a private home. We will not include patients below the age of 1 because cardiac arrests in neonates and infants are substantially different compared to cardiac arrests in children and adults. Shockable rhythms are very rare in neonates and infants. Cardiac arrest in a private home will be defined based on the CARES variable “18. Location Type” = “Home/Residence”. This will not include nursing homes. We will also not include cardiac arrests that are witnessed by 911-responders as the application of AEDs are not relevant in this population. This will be defined based on the CARES variable “19. Arrest Witness Status” = “Witnessed by 911 Responder”.

Patients will be stratified according to the initial cardiac arrest rhythm into shockable and non-shockable rhythms based on the CARES variable “29. First Arrest Rhythm of Patient”. Shockable rhythms include ventricular fibrillation, ventricular tachycardia, and “Unknown Shockable Rhythm”. Non-shockable rhythms include asystole, pulseless electrical activity, and “Unknown Unshockable Rhythm”.

### Exposure

The exposure will be application of an AED prior to EMS arrival as defined by the CARES variable “25. Was an AED Applied Prior to EMS Arrival”, irrespective of who applied the AED. The options “Yes, with defibrillation” and “Yes, without defibrillation” will be combined into one category. Given that we are combining these categories, we are technically estimating the effect of AED application, as compared to AED defibrillation. However, we assume that most patients with a shockable rhythm and AED application will also receive defibrillation by the AED.

### Outcomes

The primary outcome will be survival to hospital discharge as defined by the CARES variable “49. Hospital Outcomes” and the option “Discharged Alive”.

Secondary outcomes will include return of spontaneous circulation, admission to hospital, and favorable neurological outcome at hospital discharge. Return of spontaneous circulation is defined as either return of spontaneous circulation for at least 20 minutes or spontaneous circulation present at the end of EMS care (CARES variable #30). Admission to the hospital is defined by the CARES variable “47. ER Outcome” = “Admitted to hospital”. Favorable neurological outcome will be defined as survival with a Cerebral Performance Category (CPC) of 1 (mild or no neurological/functional deficit) or 2 (moderate cerebral disability but sufficient cerebral function for independent activities of daily life) at hospital discharge (CARES variable #51). The CPC score is determined by data abstractors reviewing the medical record.

### Additional data

Additional variables not mentioned above are listed below. They are divided based on CARES required data elements, CARES supplemental data elements, and census-level data. For additional information, see the CARES website.[3]

#### Required data elements

| Variable | Categories | Notes |
| --- | --- | --- |
| Age | Continuous variable in years | - |
| Gender | Male<br>Female<br>Non-binary*<br>Female-to-male, transgender male*<br>Male-to-female, transgender female* | Due to expected low sample sizes, “Non-binary”, “Female-to-male, transgender male”, and “Male-to-female, transgender female” will be combined into an “Other” category. |
| Race/ethnicity | American-Indian/Alaska Native<br>Asian<br>Black/African American<br>Hispanic/Latino<br>Native Hawaiian/Pacific Islander<br>White<br>Unknown<br>Multi-racial* | Due to expected low samples sizes, “American-Indian/Alaska Native”, “Native Hawaiian/Pacific Islander” and “Multi-racial” will be combined into an “Other” category. |
| Day of the week | Weekday (Monday to Friday)<br>Weekend (Saturday to Sunday) | Derived from “14. Date of the Arrest”. |
| Time of the year | Winter (December to February)<br>Spring (March to May)<br>Summer (June to August)<br>Fall (September to November) | Derived from “14. Date of the Arrest”. |
| Year | Continuous variable | Derived from “14. Date of the Arrest”. |
| Witnessed status | Unwitnessed<br>Witnessed by bystander | Patients with “Witnessed by 911 responder” are excluded. |
| Presumed etiology | Presumed cardiac etiology<br>Respiratory/asphyxia<br>Drowning/submersion<br>Electrocution<br>Exsanguination/hemorrhage<br>Drug overdose<br>Other | Patients with “Trauma” are excluded.<br>Due to expected low sample sizes, “Drowning/Submersion”, “Electrocution”, “Exsanguination/Hemorrhage”, and “Other” will be combined into a “Other” category. |
| Who initiated CPR | Not applicable<br>Bystander<br>Family member<br>Healthcare provider (non-911 responder)<br>First responder | “Not applicable” is only meaningful when an AED terminated the cardiac arrest prior to EMS arrival and those that applied the AED did not perform CPR (presumably a rare event). To avoid |
|  | EMS responder | collinearity between this variable and our exposure, “Not applicable” will be combined with “EMS responder”. |
| Who First Applied the AED | Bystander<br>Family Member<br>Healthcare provider (non-911 Responder)<br>Law Enforcement First Responder<br>Non-Law Enforcement First Responder | This variable will only be used for descriptive purposes in those who had an AED applied, i.e., the variable is not used in any of the analyses. |

\* Options added to CARES in 2021

#### Supplemental data elements

Variables from the supplemental data set will be used for subgroup and sensitivity analyses. The only variables used from the supplemental dataset are those related to EMS response time and prior medical history (CARES variable “12.Medical History”). “60. Time call received at dispatch center” and “68. Time ambulance arrived at scene” will be used to calculate EMS response time in minutes consistent with the approach used in the CARES registry.[4] This variable will only be used for assessing heterogeneity of treatment effect (see below).

#### Census-level data

Once a year, CARES data is geocoded to a United States census tract by the Centers for Disease Control and Prevention on the basis of the address of the cardiac arrest. Census-level variables are then obtained from the American Community Survey. Census variables are only available from 2013 to 2021 and will therefore only be used in sensitivity analyses. The following census variables (all continuous) will be used: median household income, median age, proportion of white people, proportion with a high school degree or higher (25 years or older), proportion that are unemployed (16 years or older), proportion below poverty level, and average household size.

## STATISTICAL METHODS

### Methodological assumptions

We are interested in estimating the effect of AED application in patients with an initial shockable rhythm. Consistent with our previous study, we will assume that AEDs have no (negative or positive) effect in patients with a non-shockable rhythm.[1] Theoretically, there could be potential detrimental effects of AED application in patients with a non-shockable rhythm such as interruptions in CPR.[5-7] Contrary, AED application could also be beneficial in patients with a non-shockable rhythm, for example through voice prompts to guide cardiopulmonary resuscitation (CPR).[8, 9] Although this has not been tested in large studies, small studies have found no beneficial effect of AED feedback.[10, 11] In our previous study of AED application for cardiac arrests in public locations, we found no meaningful association between AED application and outcomes in those with a non-shockable rhythm.[1] With these considerations in mind, we consider it reasonable to assume that there is no effect on survival of AED application in patients with a non-shockable rhythm.

Simplified Directed Acyclic Graphs (DAGs) for the relationship between AED application and survival for those with a shockable and non-shockable rhythm are provided below:

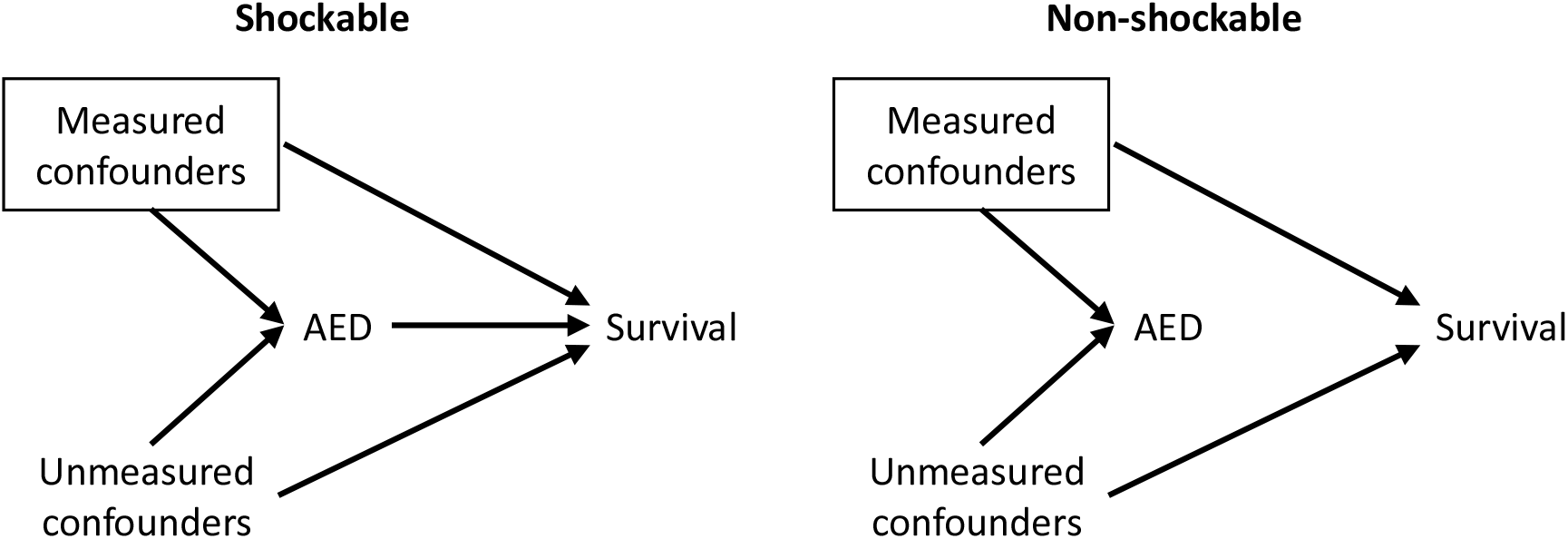

The square around “measured confounders” indicate that we can adjust for these in our analyses. However, there might also be unmeasured confounders (or residual confounding related to measured variables). These could include bystander characteristics such as bystander cardiopulmonary resuscitation training, which could lead to application of an AED and increase survival (irrespective of AED use) through higher quality chest compressions. Such bystander characteristics are rarely captured in cardiac arrest registries.

Given that the underlying rhythm is unknown prior to AED application, we will assume that there is no relationship between these two variables (see Appendix 1 for some additional considerations). We therefore consider it likely that the unmeasured confounders are similar for those with a shockable and non-shockable rhythm, i.e., the confounders that influence both AED application and survival are the same irrespective of the underlying rhythm. Given these assumptions, cardiac arrests with a non-shockable rhythm can function as “negative controls”.[12] Given our assumption of no effect of AEDs in those with a non-shockable rhythm, any association between AED application and outcomes in this patient group will be due to confounding (or theoretically other biases). Assuming that this confounding is the same in those with a shockable rhythm, we can “deduct” this confounding from the association between AED application and outcomes in those with a shockable rhythm. Under the mentioned assumptions, this will give us an unbiased estimate of the causal effect of AED application in those with a shockable rhythm. Contrary to many other statistical methods, this approach accounts for both measured and unmeasured confounding.

Given that our outcomes are binary, effects can be measured on either relative (e.g., odds ratios, risk ratios) or absolute (e.g., risk difference) scales. If baseline outcome proportions are different in two groups and there is an effect, conclusions about effect measure modification will depend on the scale being used.[13] This is relevant here, because survival is substantially lower in patients with a non-shockable rhythm. For example, if survival is 5% with AED application and 4% without AED application in those with a non-shockable rhythm, and this is assumed to be due to confounding, the “effect” of confounding can be measured as 1% (risk difference) or 1.25 (risk ratio). If survival is 30% and 20% in those with a shockable rhythm, the “deduction” of confounding can either be done on the risk difference scale, i.e., 10% - 1% = 9% or on the risk ratio scale, i.e., 1.50/1.25 = 1.20, potentially resulting in different conclusions.

It is unclear whether a measure of confounding is transferable on the relative or the absolute scale. However, meta-epidemiological studies suggest that relative effects (e.g., risk ratios and odds ratios) are generally more constant across baseline risks as compared to absolute effects.[14-16] This suggests that any confounding is also likely to be more transferable on the relative scale. Our primary analysis will therefore be on the relative scale using risk ratios. From a theoretical point of view[17], odds ratios might be more suitable as calculated probabilities are constrained between 0% and 100%. However, as we are not expecting very small (i.e., close to 0%) or very high (i.e., close to 100%) probabilities, this is likely of limited importance. Considering that odds ratios are likely harder to interpret for the general reader[18], we have decided to focus on risk ratios. Additional analyses will be conducted on the risk difference and odds ratio scales (see below).

There is one important limitation to this “difference-in-difference” approach as described below. Given that we are estimating an interaction, there will be increased uncertainty in our estimate (i.e., wider confidence intervals). This is a consequence of accounting for both the uncertainty in the estimate for the shockable group and the uncertainty in the estimate for the non-shockable group (i.e., the uncertainty in the “effect” of confounding). We consider this bias-variance tradeoff (i.e., lower bias, more variance) reasonable.

### Statistical analysis

Descriptive data will be reported as medians and 1^st^ and 3^rd^ quartiles for continuous variables and as counts and proportions for categorical variables. A template of a Table 1 is provided in Appendix 2.

Differences in outcomes between groups will be presented as risk ratios.[18] Risk ratios will be estimated using generalized linear models with a binomial distribution and log link function.[19] If this model fails to converge, a modified Poisson regression model will be used instead (i.e., Poisson distribution and log link function with robust standard errors).[19-21]

All models will be adjusted for age, race/ethnicity, day of the week, time of the year, year, witnessed status, presumed etiology, and who initiated cardiopulmonary resuscitation. Age and year will be included as continuous variables with linear and quadratic terms[22], while the remaining variables will be included as categorical variables. We will also include an interaction between sex and age.[23] These variables were chosen because they were either assumed to be confounders of the exposure-outcome relationship or predictors of the outcome, and not affected by the exposure.[24]

The causal effect in those with a shockable rhythm will be estimated using a “difference-in-difference” approach by including an interaction term between AED application and the initial rhythm in the model. The primary analysis will be done on the risk ratio scale given the assumptions listed above.[25, 26] Results will be presented in a table (see Appendix 2 for a template).

### Clustering

The CARES registry does not collect data on whether a patient is included more than once in the registry (i.e., whether the same patient has multiple cardiac arrests). It is therefore not possible to account for within-patient correlation. Given that repeat cardiac arrests are relatively uncommon[27], we believe this is of minor importance.

Potential between-patient clustering at the EMS agency level[28] will be accounted for using generalized estimating equations with an exchangeable correlation structure meaning that we will be estimating population-averaged treatment effects.[29]

### Missing data

We expect missing data on some variables especially race, census characteristics, and neurological outcome. There might also be limited missing data on other variables.

Missing or unknown data for race are primarily a result of certain communities deciding not to provide these data rather than a result of incomplete data entry.[30] Those with missing or unknown data on race in CARES have a relatively similar distribution of race compared to those with data on race as determined by linkage to other registries.[31, 32] Missing data for neighborhood characteristics are due to inability to link a cardiac arrest to a census tract or missing data in the census files. Missing data for other variables, including the exposure and outcomes, are missing due to lack of relevant data in the medical records.

The overall pattern of missingness is assumed to be arbitrary (i.e., non-monotone) and “missing at random”.[33, 34] Considering the mechanism for missingness described above, we consider these assumptions reasonable.

Missing data will be reported for each variable and a comparison of those with any missing data and those with no missing data will be provided in a supplemental table.[35]

Missing values will be imputed using multiple imputations by chained equations.[36-39] Binary variables will be imputed using logistic regression models and nominal variables with discriminant analysis.[40] Continuous variables will be imputed using linear regression unless data is severely non-normally distributed in which case other distributions will be considered. For each dataset, 20 burn-in iterations will be used before the imputation.[36]

The imputation model will include all available variables including the exposure and the outcomes.[35, 37] The multiple imputation will be stratified according to the initial rhythm.[41] Continuous variables will be included as linear and quadratic terms and an interaction between sex and age will be included.[23] A total of 100 data sets will be created.[37, 42, 43] Results from the 100 data sets will be combined using Rubin’s rule.

### Heterogeneity of treatment effect

We will assess heterogeneity of treatment effect for age, gender, and EMS response time by including an interaction term between AED application and these variables in the models. For all these analyses, we will only evaluate the shockable group (i.e., we will not perform the “difference-in-difference” analysis described above). Age and EMS response time will be treated as continuous variables, and we will consider both linear and quadratic terms. Results for the continuous variables will be presented graphically. Gender will be classified as male or female, and those categorized as transgender male, transgender female, or non-binary will be excluded for this analysis given the presumed low sample sizes in these groups.

### Additional analysis

We will conduct several additional analyses:

1. Some patients with a shockable rhythm will have an AED applied but not receive defibrillation. While these patients are categorized as “AED applied” in our analysis, they will not benefit from the AED application and results from our analysis will therefore likely be closer to the null compared to an analysis examining AED defibrillation. We will conduct an analysis where the exposure is AED defibrillation. This analysis can only be performed in those with a shockable rhythm and the difference-in-difference analysis will therefore not be performed.
2. Census-level variables are potentially confounders. We will therefore conduct analyses where these variables are adjusted for. All variables will be included in the model as continuous variables using both linear and quadratic terms. This analysis will be restricted to cardiac arrests from 2013 to 2021 and cardiac arrests that are matched to a census tract (i.e., cardiac arrest without missing data on the census variables).
3. Medical history is a potential confounder. However, this variable is only part of the supplemental data elements and is therefore missing for a large proportion of the patients. We will conduct analyses where medical history is included in the model. Missing data (including “unknown” for medical history) will be imputed as described above.
4. Although our primary results will be presented on the relative risk scale, we will conduct similar analyses on the absolute risk difference and the odds ratio scales. Absolute risk differences will be estimated using generalized linear models with a binomial distribution and an identity link function. If this model fails to converge, a modified Poisson approach will be used.[19] Odds ratios will be estimated from generalized linear models with a binomial distribution and a logit link function.

### P values and confidence intervals

Since our primary goal is estimation and not null-hypothesis significance testing, P values will not be reported. All confidence intervals will have 95% coverage and will not be adjusted for multiplicity.

### Sample size considerations

Given that we are using already collected data, our sample size is fixed. We have therefore not performed any formal sample size calculation. In the below, we provide some considerations regarding the sample size and power. Aggregate baseline data (not outcomes) were provided by the CARES registry for these calculations prior to receipt of the full dataset by the investigators.

Approximately 550,000 patients in the CARES registry meets the inclusion criteria for the current study. Of these, approximately 1,200 (0.2%) had an AED applied. We anticipate that about 20% of the overall group will have a shockable rhythm (i.e., 110,000) and 80% (i.e., 440,000) will have a non-shockable rhythm.[44, 45] We will assume that AED application is similar in those with a shockable and non-shockable rhythm, i.e., approximately 240 and 960 patients, respectively. In the shockable group, we anticipate a relative effect size like that for public cardiac arrest (i.e., a risk ratio of about 1.5).[1] We expect that survival in the non-AED group will be approximately 20% and therefore 30% in the AED group. We assume survival of 5% in the non-shockable group with no effect of AED application.

Using these estimates, we simulated 10,000 data sets with outcomes simulated using Bernoulli distributions and the remaining quantities being fixed. For each dataset, we then calculated the “difference-in-difference” as described above estimating the risk ratio with 95% confidence intervals for the effect of AED application on survival in those with a shockable rhythm. The SAS code is provided in Appendix 3. The distributions for the point estimate and the upper and lower bound of the 95% confidence interval are provided below.

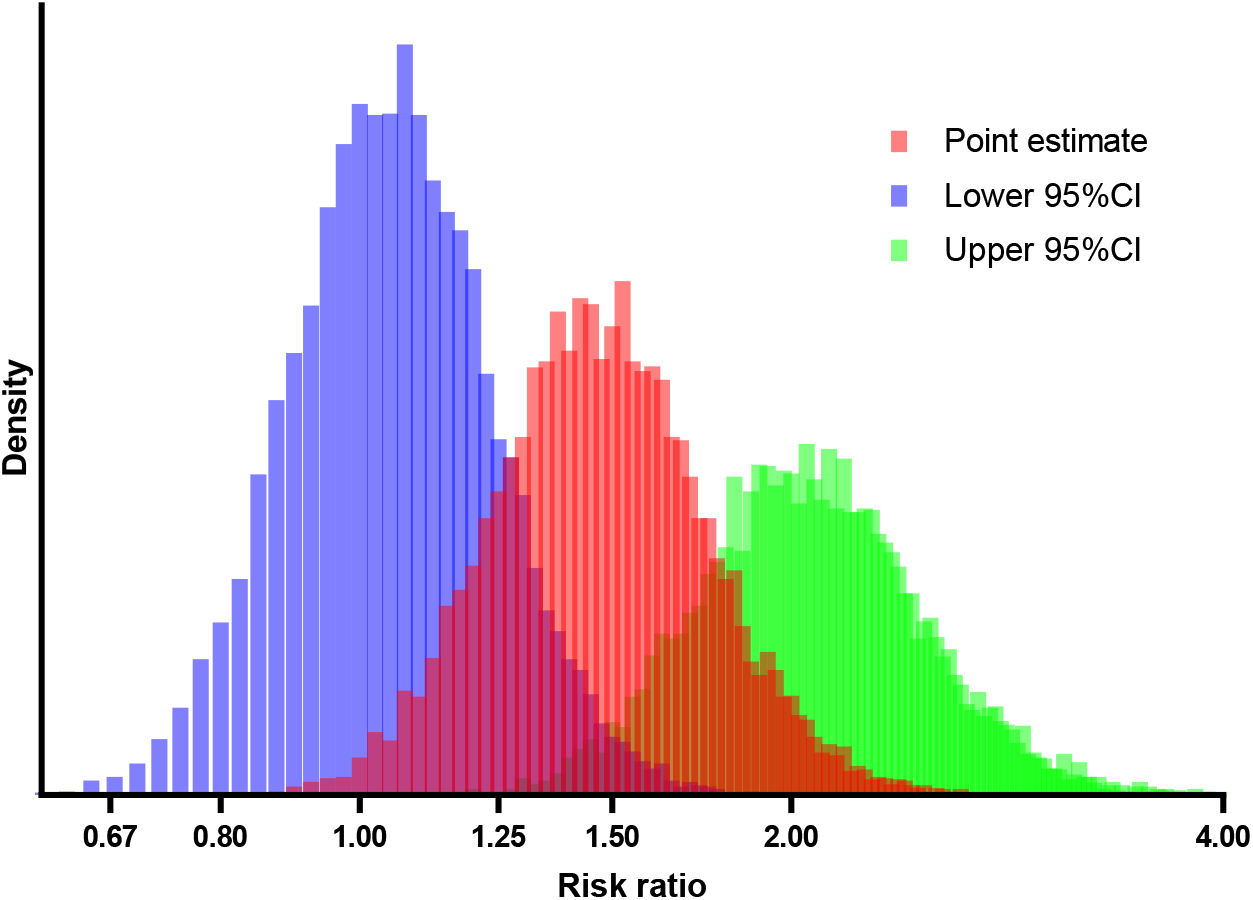

The proportion of datasets with the lower bound of the 95% confidence interval above 1 was 65%, indicating that power at an alpha of 5% is 65%. We will likely have higher power, as we are adjusting for a number of strong predictors of the outcome.[46]

We note that the CARES registry is one of the largest out-of-hospital cardiac arrest registries in the world[47], and the only large one, we are aware of, that have the available data (specifically the application of an AED irrespective of defibrillation) for the current analysis.

### Software

All analyses will be performed using SAS statistical software, version 9.4 or higher (SAS Institute Inc), or R software, version 4.2.2 or higher (R Foundation for Statistical Computing).

## DISSEMINATION, DATA SHARING AND ETHICS

### Dissemination

Study results will be published irrespective of the findings. The manuscript will adhere to STROBE reporting guidelines[48] and will include the statistical code as a supplement. Authorship will follow authorship guidelines from the International Committee of Medical Journal Editors.[49]

### Data sharing

Information on access to the CARES registry, including information regarding the application process, can be found on the CARES website.[3]

### Ethical approval

The Institutional Review Board at Emory University have determined that the CARES registry does not require Institutional Review Board approval as it is a public health surveillance registry and quality improvement program.[50] Retrospective, register-based research does not require ethical committee review or approval in Denmark, where the investigators are located.[51]

## CONFLICTS

The authors have no relevant conflicts of interest.

## Data Availability

NA

## ACKNOWLEDGEMENTS

We would like to thank Rabab Al-Araji, M.P.H., senior epidemiologist at Emory University, for providing aggregate data from the CARES registry and for answering data-related questions.

### APPENDIX 1. Additional methodological considerations

In our approach, we assume no relationship between the initial rhythm and AED application. There are two caveats to this assumption illustrated in the below DAG.

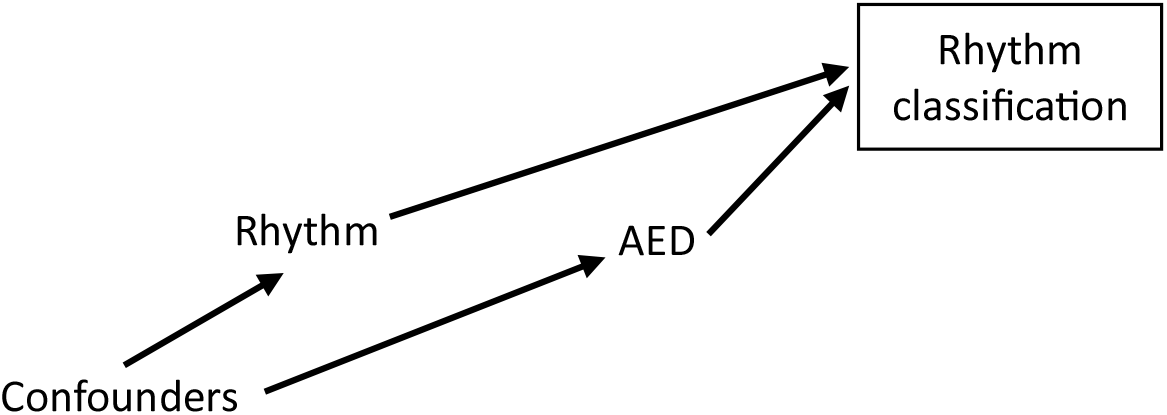

First, there might be variables that are associated with both the initial rhythm and the application of an AED (i.e., confounders), which would introduce a (non-causal) relationship between the initial rhythm and AED application. Consider, for example, socioeconomic status, where people with a higher socioeconomic status might have a different disease profile that would lead to a different initial rhythm in the case of a cardiac arrest. Higher socioeconomic status might also increase the availability and application of AEDs.[1] We consider this issue of limited importance as we are conditioning on a number of measured confounders (including measures of socioeconomic status in our additional analyses). Any residual relationship between rhythm and AED application is likely going to be minimal.

Second, the actual initial rhythm is unknown and the rhythm we use is the first one recorded. Given that AEDs are applied earlier than manual defibrillations, the application of AEDs might increase the proportion of patients classified as having a shockable rhythm, as these patients might deteriorate to a non-shockable rhythm before a manual rhythm is performed.[52, 53] By conditioning on rhythm classification (illustrated by the square in the DAG), a non-causal relationship is created between rhythm and AED application due to collider bias.[54] This could lead to a biased relationship between AED application and survival, even if there is no causal relationship, as illustrated in the below graph.

However, again, we are conditioning on many variables that are associated with both the initial rhythm and survival, and we therefore believe this bias is likely to be small. This is also supported by prior empirical data, where the proportion of cardiac arrests with a shockable rhythm was only marginally higher when an AED was applied.[2] For additional discussion and references, see page 10-11 of the supplemental material in Andersen et al.[2] It is difficult to predict the direction of any such bias if present. If we assume that those who transition from a shockable to a non-shockable rhythm (potentially classified as shockable only when an AED was applied) have worse outcomes compared to those who do not transition (i.e., stay in a shockable rhythm), but have better outcomes than those who have a non-shockable rhythm throughout[55], the bias will likely favor the no AED group in both the shockable group and the non-shockable group. However, we assume that any such bias will be minimal.

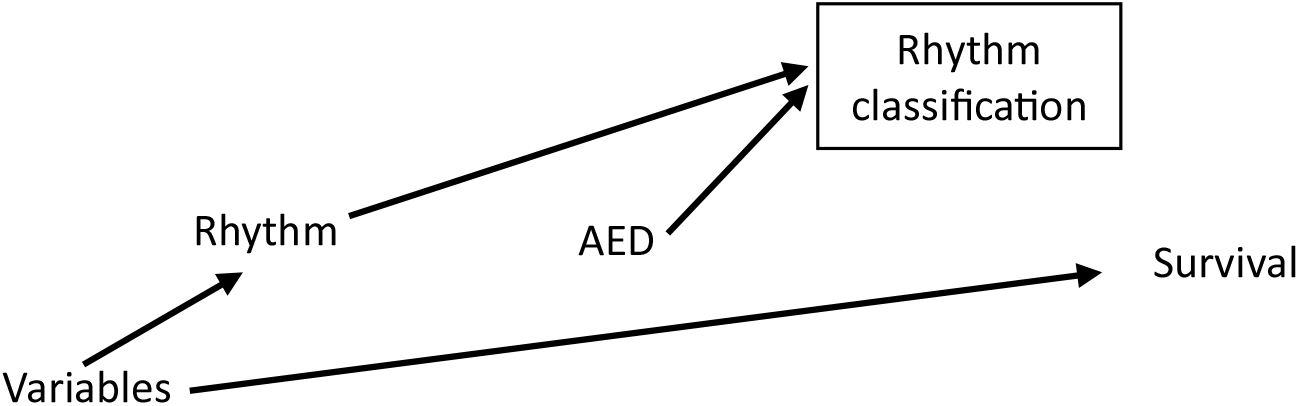

The classified etiology of the cardiac arrest (as a surrogate of the actual etiology) might be causally affected by the rhythm classification (e.g., a shockable rhythm might lead someone to classify the cardiac arrest as having a presumed cardiac etiology). Conditioning on classified etiology can therefore also introduce collider bias. However, etiology is also a potential confounder and considering the above, we will therefore adjust for it in the analyses.

### APPENDIX 2. Table templates

**Table 1.**
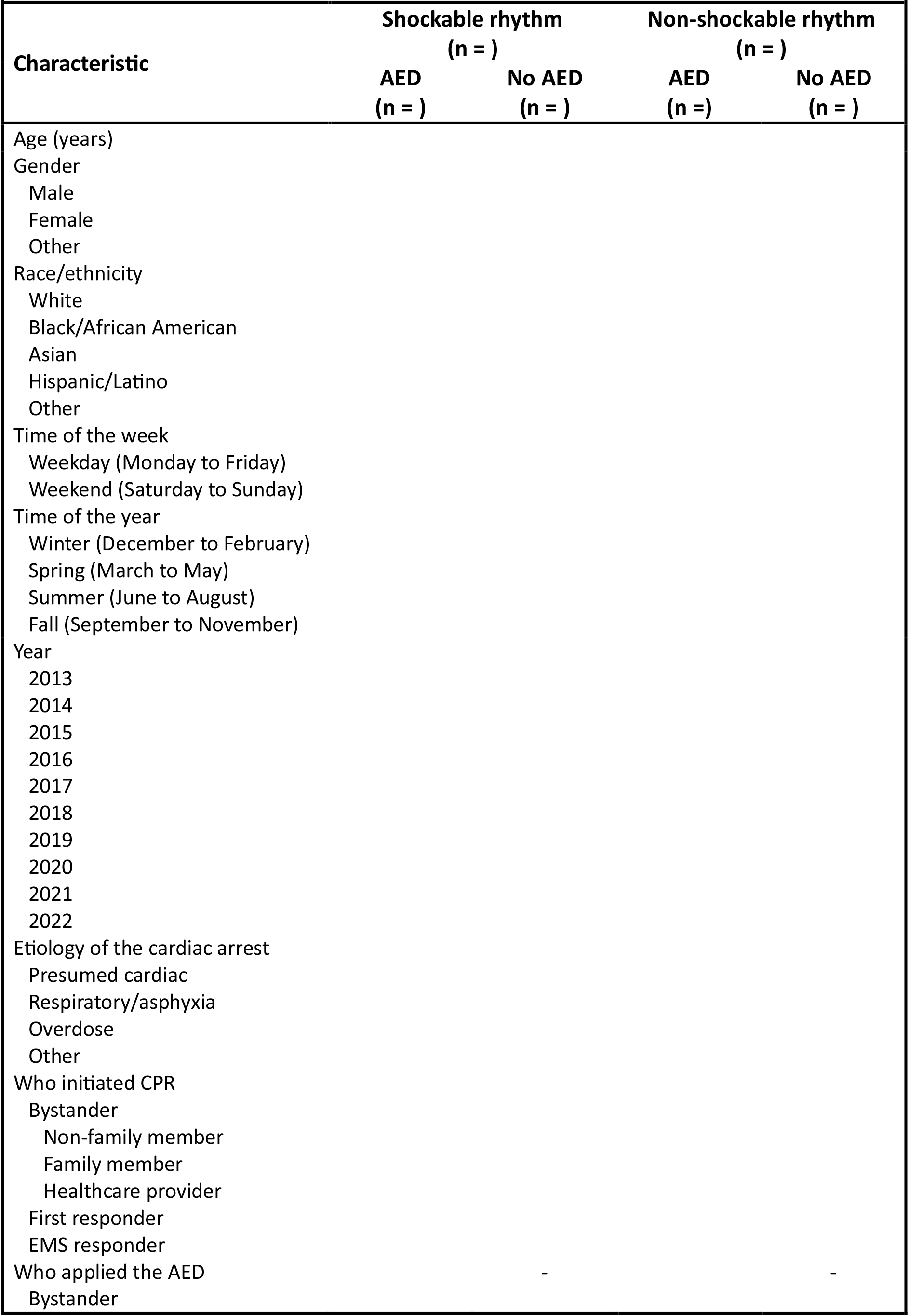
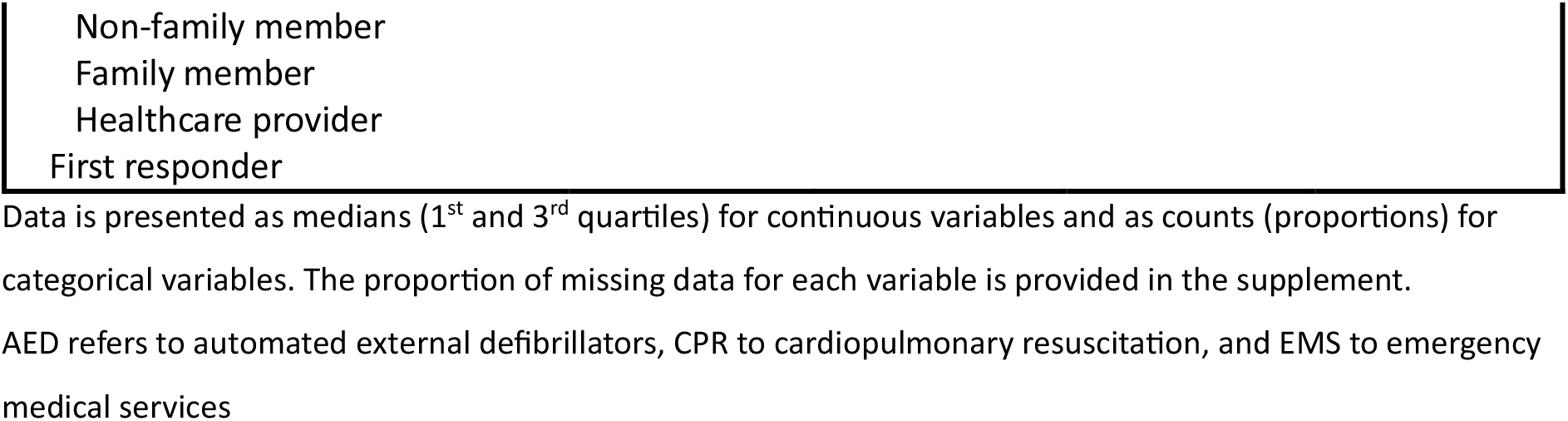
Baseline characteristics according to initial rhythm and AED application.

**Table 2.** AED application and outcomes.

| Table 2. AED application and outcomes |  |  |  |
| --- | --- | --- | --- |
|  | No AED<br>- No. (%) | AED<br>- No. (%) | Risk ratio<br>(95% CI) |
| **Shockable rhythms (n = )** |  |  |  |
| Return of spontaneous circulation |  |  |  |
| Admission to hospital |  |  |  |
| Survival to hospital discharge |  |  |  |
| Favorable functional outcome |  |  |  |
| **Non-shockable rhythms (n = )** |  |  |  |
| Return of spontaneous circulation |  |  |  |
| Admission to hospital |  |  |  |
| Survival to hospital discharge |  |  |  |
| Favorable functional outcome |  |  |  |
| **Difference-in-difference (n = )** |  |  |  |
| Return of spontaneous circulation | - | - |  |
| Admission to hospital | - | - |  |
| Survival to hospital discharge | - | - |  |
| Favorable functional outcome | - | - |  |

AED refers to automated external defibrillators

### APPENDIX 3. SAS code for simulations

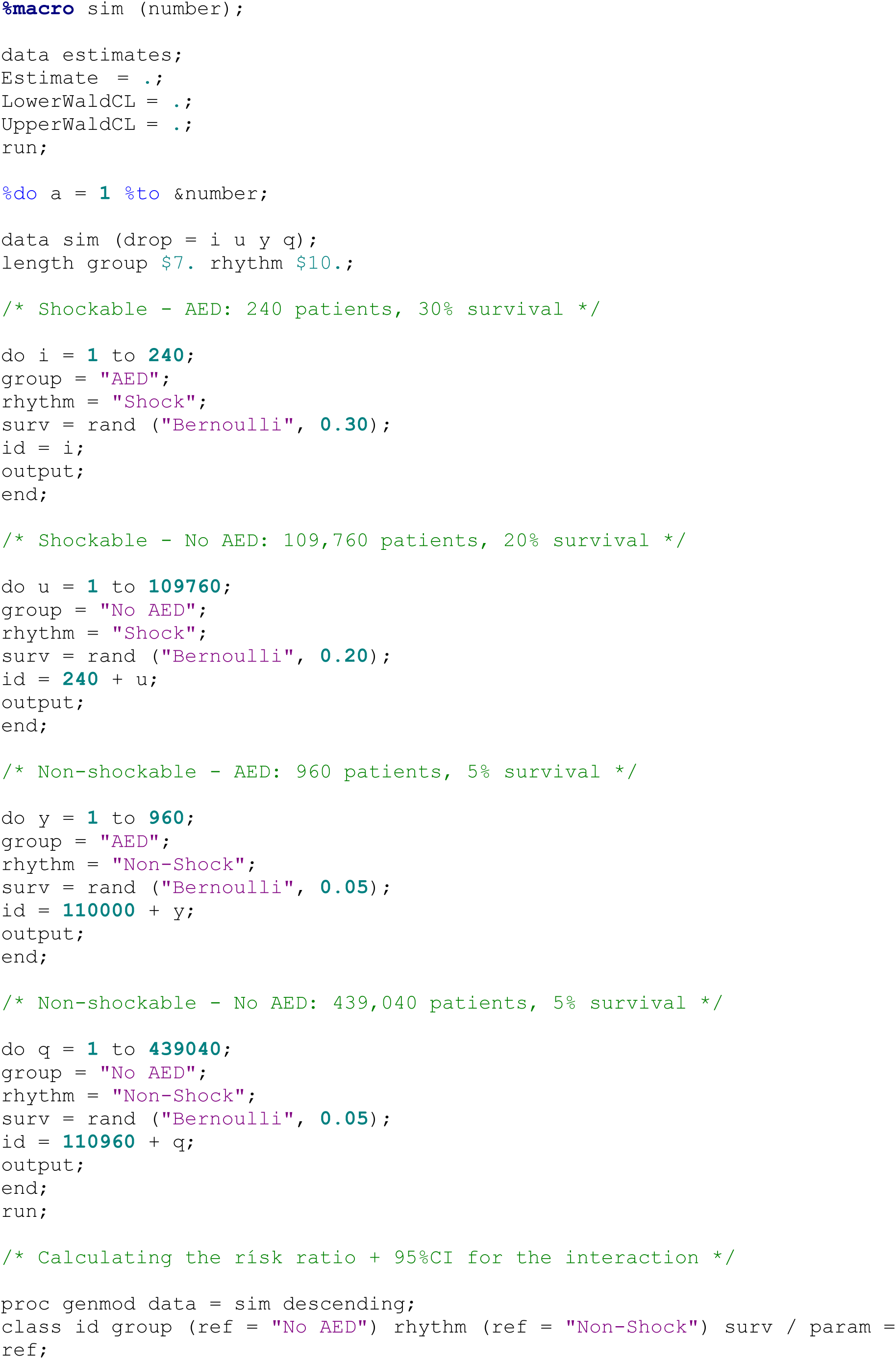
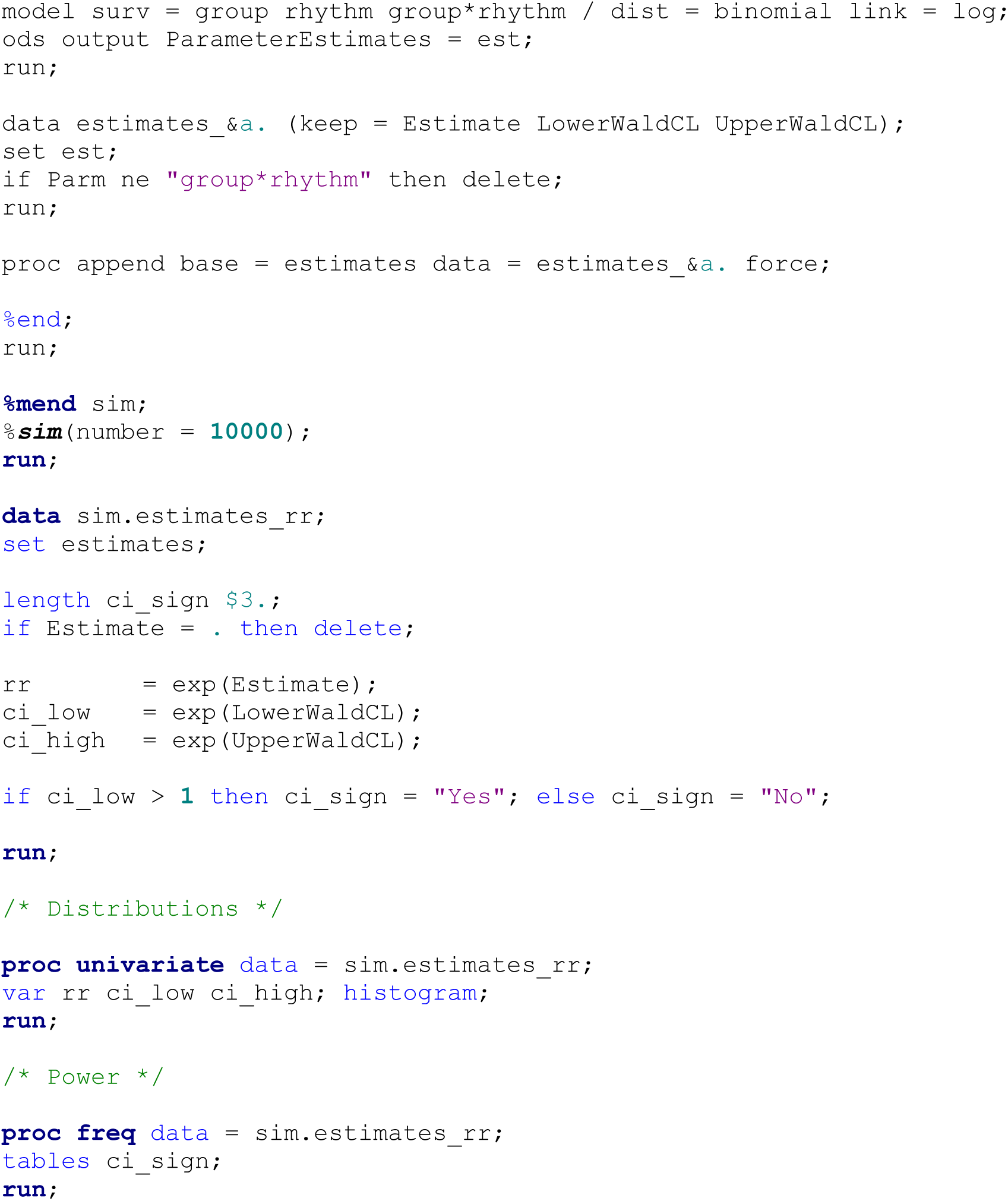

